# Somatically mutated genes in fatty liver disease have minimal influence on germline risk

**DOI:** 10.1101/2021.10.18.21265159

**Authors:** Jake P. Mann, Matthew Hoare

## Abstract

**Background:** Understanding the genetics of liver disease has the potential to facilitate clinical risk stratification. We recently identified six genes and one lncRNA enriched for acquired somatic mutations in patients with NAFLD and alcohol-related liver disease. We hypothesised that germline variation in these genes would be associated with risk of liver disease development and contribute to prognostication.

**Methods:** Genome-wide association study (GWAS) summary statistics were extracted from seven studies (>1.7 million participants) for variants near *ACVR2A, ALB, CIDEB, FOXO1, GPAM, NEAT1* and *TNRC6B* for: aminotransferases, liver fat, HbA1c, diagnosis of NAFLD, ARLD, and cirrhosis. Findings were replicated using GWAS data from multiple independent cohorts. A phenome-wide association study was performed to examine for related metabolic traits, using both common and rare variants, including gene-burden testing.

**Results:** There was no evidence of association between rare germline variants or SNPs near five genes (*ACVR2A, ALB, CIDEB, FOXO1*, and *TNRC6B*) and risk or severity of liver disease. Variants in *GPAM* were associated with liver fat (p=3.6×10^-13^), ALT (p=2.8×10^-39^), and serum lipid concentrations. Variants in *NEAT1* demonstrated borderline significant associations with ALT (p=1.9×10^-11^) and HbA1c, but not with liver fat, as well as influencing waist-to-hip ratio, adjusted for BMI.

**Conclusions:** Despite strong selective advantage to acquire somatic mutations at these loci, there was no evidence of an association between germline variation and markers of liver disease, except in *GPAM*. Polygenic risk scores based on germline variation alone will not capture prognostic data from genes affected by somatic mutations.

## Introduction

Non-alcoholic fatty liver disease affects around 25% of the worldwide population and is emerging as the fastest growing form of liver disease in developed countries [1,2]. It encompasses a spectrum of disease from simple hepatic steatosis, through non-alcoholic steatohepatitis (NASH) to cirrhosis, with the attendant risks of liver failure and hepatocellular carcinoma (HCC). Given the number of patients at risk of development of NAFLD and subsequent health problems, there is an urgent need to stratify patients based upon long-term risks, so that even countries with well-resourced healthcare schemes can cope with the patient numbers predicted to develop end-stage liver disease over the next few decades.

At present, there are a number of clinically-deployed non-invasive tests, including elastography and serum biomarkers that can predict levels of hepatic fibrosis and long-term risk in NAFLD. Significant interest has been generated in the potential of germline genetic variation in the pathogenesis and prognostication of NAFLD[3]. Single-nucleotide polymorphisms (SNPs) near several genes, including *PNPLA3[4], HSD17B13[5]*, and *TM6SF2[6]* have been shown to be associated with long-term risk of several disease-related outcomes in NAFLD[7], as well as in alcohol-related liver disease (ARLD)[8], through genome-wide association studies (GWAS). These variants are common, but associated with small effect sizes, when studied in isolation. This has suggested the potential for combining multiple genotypes into a polygenic risk score: the analysis of several SNPs in patients at baseline as a predictive tool for long-term outcomes in both NAFLD[9] and ARLD[8].

Similarly, other studies have focussed on rare pathogenic germline variants, such as in *APOB* the gene that encodes apolipoprotein B, that lead to NAFLD with high penetrance[10]. Although these cases can inform us about underlying disease pathogenesis, their low frequency suggests that they are unlikely to be informative in the stratification of most patients within the general NAFLD population.

We have recently identified recurrent somatic mutations within the liver of patients with NAFLD and ARLD[11]. Recurrent non-synonymous mutations were seen in 6 coding genes: *ACVR2A, ALB, CIDEB, FOXO1, GPAM, TNRC6B*, plus the long non-coding (lnc)RNA *NEAT1*. These mutations were found in multiple hepatocyte clones throughout the liver, with evidence of convergent evolution: the identification of independent clones with mutations in the same genes, suggesting strong selection pressure to acquire these variants in the context of fatty liver disease. Amongst these genes, 3 are involved in lipid metabolism suggesting a potential functional role in disease pathogenesis: *CIDEB*, mediates fusion and cargo transfer of cytoplasmic lipid droplets[12]; *FOXO1*, the main transcription factor downstream of insulin[13], but also a major regulator of lipid metabolism[14] and; *GPAM*, encoding GPAT1, the enzyme catalyzing the initial step in triglyceride synthesis[15]. Through functional validation of the hot-spot mutations in *FOXO1*, we identified that this impacted upon the response to insulin, glycolysis and lipid metabolism. Although promising for improving the mechanistic understanding of disease pathogenesis, the prognostic or therapeutic role that these acquired mutations could play in NAFLD and ARLD remains unknown.

As there was strong selective pressure to acquire these mutations, we were interested to explore whether common or rare germline variation near these recurrently mutated genes might be associated with liver-related outcomes in NAFLD. We hypothesised that germline variation providing weaker modulation of disease phenotypes would exist and might improve the performance of polygenic risk scores being developed for prognostication in NAFLD and ARLD.

Amongst these genes, germline coding variants *GPAM* have been associated with serum ALT levels in an exome-wide association study of the UK Biobank cohort[16]. Further study found that these variants were also associated with hepatic fat content and histological markers of liver damage in independent NAFLD cohorts. Other studies using GWAS have replicated these findings in further cohorts[17–19], where rs10787429 in *GPAM* was associated with elevated ALT in both NAFLD and ARLD. Here we use data from multiple GWAS to investigate whether similar associations are observed for the other 5 recurrently mutated genes and one recurrently mutated locus.

## Methods

### Identification of genes enriched for somatic mutations in ARLD and NAFLD

As described in detail elsewhere[11], whole genome sequencing was performed on diseased, non-malignant hepatic tissue from patients with NAFLD and ARLD undergoing tumour resection or liver transplant. The dN/dScv method[20] was used to identify six coding genes with higher numbers of nonsynonymous mutations relative than expected.

### Annotation of somatic mutants and genomic regions

After removal of duplicates, somatic mutations at *ACVR2A, ALB, CIDEB, FOXO1, GPAM*, and *TNRC6B*, plus one non-coding lncRNA *NEAT1[11]* were annotated with predicted consequence, impact on transcript, and prevalence within the 1000 Genomes dataset[21] (n=2,504), Exome Sequencing Project[22] (n=6,503), and gnomAD[23] (n=141,456) using the Ensembl Variant Effect Predictor[24]. For the variants that had previously been identified in any of the above population genomic sequencing datasets, we searched Phenoscanner[25] and the Common Metabolic Disease Portal[26] for any evidence of association with metabolic traits. No data was available on Phenoscanner for these ultra-rare variants. The Common Metabolic Disease Portal search yielded 96 variant-trait associations therefore, rather than apply genome-wide association significance cut-off, the critical p-value for significance for this analysis only was p<5.2×10^-4^ (i.e. 0.05 / 96).

In addition, we annotated each of the six genes with their expected and observed number of predicted loss of function (pLoF) and missense mutations from gnomAD[23]. In brief, the ratio between observed and expected pLoF mutants is an indicator of each gene’s tolerance to haploinsufficiency. Such that a gene with a low observed / expected ratio (defined as the upper 95% confidence interval <0.35) has fewer pLoF mutants than other genes, suggesting that humans are relatively intolerant of haploinsufficiency.

### Association with markers of liver disease from genome-wide association studies

Summary statistics from genome-wide association studies (GWAS) were searched for all variants within the genomic regions for the seven regions of interest (**SupTable 1**). Regions included were (GRCh37): *ACVR2A*: chr2:148,602,086-148,688,393; *ALB*: chr4:74,262,831-74,287,129; *CIDEB*: chr14:24,774,302-24,780,636; *FOXO1*: chr13:41,129,804-41,240,734; *GPAM*: chr10:113,909,624-113,975,135; *NEAT1*: chr11:65,190,245-65,213,011; *TNRC6B*: chr22:40,440,821-40,731,812.

In addition, for comparison, we extracted summary statistics for four well-validated genome-wide significance risk variants for fatty liver disease: rs738409C>G in *PNPLA3[4]*, rs58542926C>T in *TM6SF2[6]*, rs2642438A>G in *MTARC1[27]*, and rs72613567TA>T in *HSD17B13[5]*. However, for some studies data on rs72613567TA>T was not available therefore we used rs13125522A>G as a proxy, which is in strong linkage disequilibrium (R^2^=0.97) with rs72613567[28].

Summary statistics were obtained from the Pan-UK BioBank analysis[29] for alanine aminotransferase (ALT), aspartate aminotransferase (AST), glycosylated haemoglobin (HbA1c), diagnosis of NAFLD (phecode-571.5), diagnosis of ARLD (phecode-317.11), liver fibrosis (icd10-K74), cirrhosis (phecode-571), and other liver diseases (phecode-571.5).

We also obtained summary statistics for diagnosis of NAFLD from Anstee et al.[30] and MRI liver fat from Liu et al.[31], which utilises UK BioBank data. These data were accessed through GWAS Catalogue[32]. For replication of findings, we first obtained categorical data on liver-related diagnoses from the FinnGen study: NAFLD, ARLD, hepatocellular carcinoma and intrahepatic cholangiocarcinoma, and cirrhosis. For replication of observations for ALT, we obtained data from BioBank Japan[33] and Pazoki et al.[34]. For replication of findings for HbA1c, we obtained summary statistics on HbA1c from the MAGIC consortium (trans-ancestry meta-analysis by Chen et al.[35]) and BioBank Japan, plus diagnosis of type 2 diabetes mellitus (T2DM) in East Asian individuals from Spracklen et al.[36]. The total number of unique participants included from these GWAS summary statistics was 1,628,945.

All variants from the above regions were extracted, and coordinates from FinnGen were carried over from GRCh38 to GRCh37 using the Ensembl Assembly Converter. Manhattan plots were produced for each trait, illustrating only variants within the regions of interest. Significance was defined as p<5×10^-8^. Within regions that had variants with significant association, we used FIVEx to look for expression quantitative trait loci (eQTL), which extracts data from the European Bioinformatic Institute eQTL Catalogue[37].

### Gene-based phenome-wide association study for common variants

We sought to explore associations between germline variation in the coding genes and lncRNA of interest and metabolic traits using a phenome-wide association study approach. Phenoscanner[25] and the Common Metabolic Diseases Knowledge Portal[26] were searched for each of the six coding genes plus *NEAT1*. Rare variants (mean allele frequency (MAF) <0.01) were excluded and results were filtered for traits relevant to ARLD and NAFLD. Significance was defined as p<5×10^-8^. Data on eQTLs was obtained using the Qtlizer package for R[38] for all significant associations.

We also searched for any metabolite-wide associations within the regions of interest using data from Lotta et al.[39] (https://omicscience.org/apps/crossplatform/); however, we did not identify any significant (p<4.9×10^-10^, as defined by the authors) associations.

### Association between rare coding variants and traits related to NAFLD or ARLD

We next investigated whether rare coding variants individually, or combined using gene-burden analyses, were associated with markers of liver disease or related metabolic traits. We used data from https://genebass.org/ [40] and https://azphewas.com/ [41], which derive data from the UK BioBank 300k Exomes. These analyses were not available for the lncRNA *NEAT1*. Extracted associations for: individual variants and gene-burden tests for predicted loss of function (pLoF), missense, and synonymous variants. Significance, as defined by the original studies, was p<2.5×10^-8^ for GeneBass (using SKAT-O test) and p<2.0×10^-9^ (-log10(8.7)) for AZPheWAS.

### Analyses

Data were analysed using R 4.0.2[42] and the code used in analyses is available from https://doi.org/10.5281/zenodo.4656979.

## Results

We have recently identified recurrent somatic mutations in non-malignant liver tissue from individuals with ARLD and NAFLD through laser capture microdissection and whole genome sequencing[11]. Through this approach we identified six coding genes (*ACVR2A, ALB, CIDEB, FOXO1, GPAM* and *TNRC6B)* and one lncRNA *(NEAT1)* significantly enriched for acquired somatic variants. We hypothesised that given the selective advantage these variants must endow, rare pathogenic variants of these regions, associated with features of liver disease, might be identified.

### Specific somatic mutations in recurrently mutated genes are not found in the germline

We previously identified 129 unique variants across these six coding genes and one lncRNA (**Supplementary Table 2**), the majority of which were either missense (49/129, 38%) or non-coding exonic variants in *NEAT1* (49/129, 38%), and predicted to have a moderate or high impact upon protein structure (61/129, 47%, **Table 1**). These variants are extremely rare in the germline, with 118/129 (92%) having never been identified across 150,463 individuals from gnomAD, 1000G, or the Exome Sequencing Project. The most common of these eleven previously reported variants was rs368997599 G>A (p.Arg45Trp) in *CIDEB*, which was identified in 10 individuals from gnomAD in the heterozygous state, 7 of whom are of Ashkenazi Jewish ancestry. Even in this genetic ancestry p.Arg45Trp in *CIDEB* remained an ultra-rare mutation with allele frequency = 7.0×10^-4^. There was no evidence of associations between any of the previously identified variants and metabolic traits in the Common Metabolic Disease Portal (**Supplementary Table 3**).

**Table 1:** Characteristics of variants identified as somatic mutants in non-malignant liver tissue. Features of 129 unique somatic mutants across six protein-coding genes and one lncRNA (*NEAT1*). Variants were annotated using Variant Effect Predictor to derive a predicted consequence and severity of impact. Germline frequency of variants was assessed across any of gnomAD, 1000G, and ESP (n=150,463). Data on observed and expected (O/E) predicted loss of function (pLoF) and missense variants was obtained from gnomAD v2.

|  | *ACVR2A* | *ALB* | *CIDEB* | *FOXO1* | *GPAM* | *NEAT1* | *TNRC6B* | Overall |
| --- | --- | --- | --- | --- | --- | --- | --- | --- |
| **Variant consequence** |  |  |  |  |  |  |  |  |
| Stop gained | 2 | 2 | 2 | 0 | 1 | 0 | 3 | 10 (7.8%) |
| Stop lost | 0 | 0 | 1 | 0 | 0 | 0 | 0 | 1 (0.8%) |
| Missense variant | 12 | 4 | 17 | 3 | 11 | 0 | 2 | 49 (38%) |
| Missense variant (in splice region) | 0 | 1 | 1 | 0 | 1 | 0 | 1 | 4 (3.1%) |
| Synonymous variant | 0 | 5 | 1 | 0 | 0 | 0 | 1 | 7 (5.4%) |
| Non-coding variant (within lncRNA) | 0 | 0 | 0 | 0 | 0 | 49 | 0 | 49 (38%) |
| Variant upstream of lncRNA | 0 | 0 | 0 | 0 | 0 | 1 | 0 | 1 (0.8%) |
| Variant downstream of lncRNA | 0 | 0 | 0 | 0 | 0 | 8 | 0 | 8 (6.2%) |
| **Impact on protein structure** |  |  |  |  |  |  |  |  |
| HIGH | 2 | 2 | 2 | 0 | 2 | 0 | 3 | 11 (8.5%) |
| LOW | 0 | 5 | 3 | 0 | 1 | 0 | 1 | 10 (7.8%) |
| MODERATE | 12 | 5 | 17 | 3 | 10 | 0 | 3 | 50 (38.8%) |
| MODIFIER | 0 | 0 | 0 | 0 | 0 | 58 | 0 | 58 (45%) |
| **Allele frequency in germline** |  |  |  |  |  |  |  |  |
| ≤.1% | 1 | 1 | 5 | 0 | 3 | 0 | 1 | 11 (8.5%) |
| Nil | 13 | 11 | 17 | 3 | 10 | 58 | 6 | 118 (91.5%) |
| **pLoF in gnomAD** |  |  |  |  |  |  |  |  |
| Observed / expected variants, n | 1 / 28 | 7 / 34 | 18 / 12 | 0 / 17 | 23 / 48 |  | 9 / 91 |  |
| Observed / expected ratio (95% CI) | 0.04 (0.01-0.17) | 0.21 (0.12-0.39) | 1.51 (1.02-1.92) | 0 (0-0.17) | 0.48 (0.34-0.68) |  | 0.1 (0.06-0.17) |  |
| **Missense in gnomAD** |  |  |  |  |  |  |  |  |
| Observed / expected variants, n | 123 / 277 | 288 / 321 | 123 / 127 | 221 / 305 | 359 / 440 |  | 733 / 987 |  |
| Observed / expected ratio (95% CI) | 0.44 (0.38-0.52) | 0.9 (0.81-0.99) | 0.97 (0.84-1.13) | 0.72 (0.65-0.81) | 0.82 (0.72-0.89) |  | 0.74 (0.7-0.79) |  |

Whilst all of the 6 coding genes had the expected number of missense mutations, four of the six (*ACVR2A, ALB, FOXO1*, and *TNRC6B*) are under selection pressure to prevent against haploinsufficiency; in gnomAD there were significantly fewer predicted loss of function (pLoF) variants observed than expected (**Table 1**). This implies that there are a reduced number of germline variants in these genes that will cause pLoF, compared to other coding genes and potential haploinsufficiency in these genes is deleterious. The exception to this was *CIDEB*, where there was an expected number of missense and pLoF mutations, but no reports of these rare variants associated with liver phenotypes. Overall, the specific somatic mutations that we had previously identified are very rare in the germline, due to negative selection pressure and not known to be associated with the development of liver disease.

### Among recurrently mutated genes only germline exonic variation in *GPAM* is associated with liver phenotypes

Given the rarity (or absence) of these specific 129 variants in the germline, we investigated whether other rare variants at these loci were associated with liver disease (or related metabolic traits). We used data from https://genebass.org/ and https://azphewas.com/ (n=281,852 from UK BioBank), which tests whether exonic germline variants either individually, or cumulatively using a gene-based burden method, demonstrated an association with phenotypes.

Analysis of individual coding variants found that p.Ile43Val in *GPAM* was associated with differences in serum lipids and the synonymous mutation 10-112157327-T-A (p.Pro681Pro) in *GPAM* was associated with ALT (**Table 2**). Exonic variants in other genes were associated with related metabolic traits, but not with liver disease phenotypes (**Table 2 & Supplementary Table 4**). Using gene-based burden testing, which adds together all variants within a single category (e.g. pLoF or missense), variants in *ALB* demonstrated associations with serum lipids, but no other markers of liver disease. Burden testing for pLoF variants in *TNRC6B* demonstrated a significant association with alcohol consumption habits, but no other markers of alcohol-related disease (**Supplementary Table 4**). Therefore, amongst the 6 recurrently mutated genes only rare germline coding variants in *GPAM* have been identified to be associated with serum lipid levels, rather than liver phenotypes.

**Table 2:**
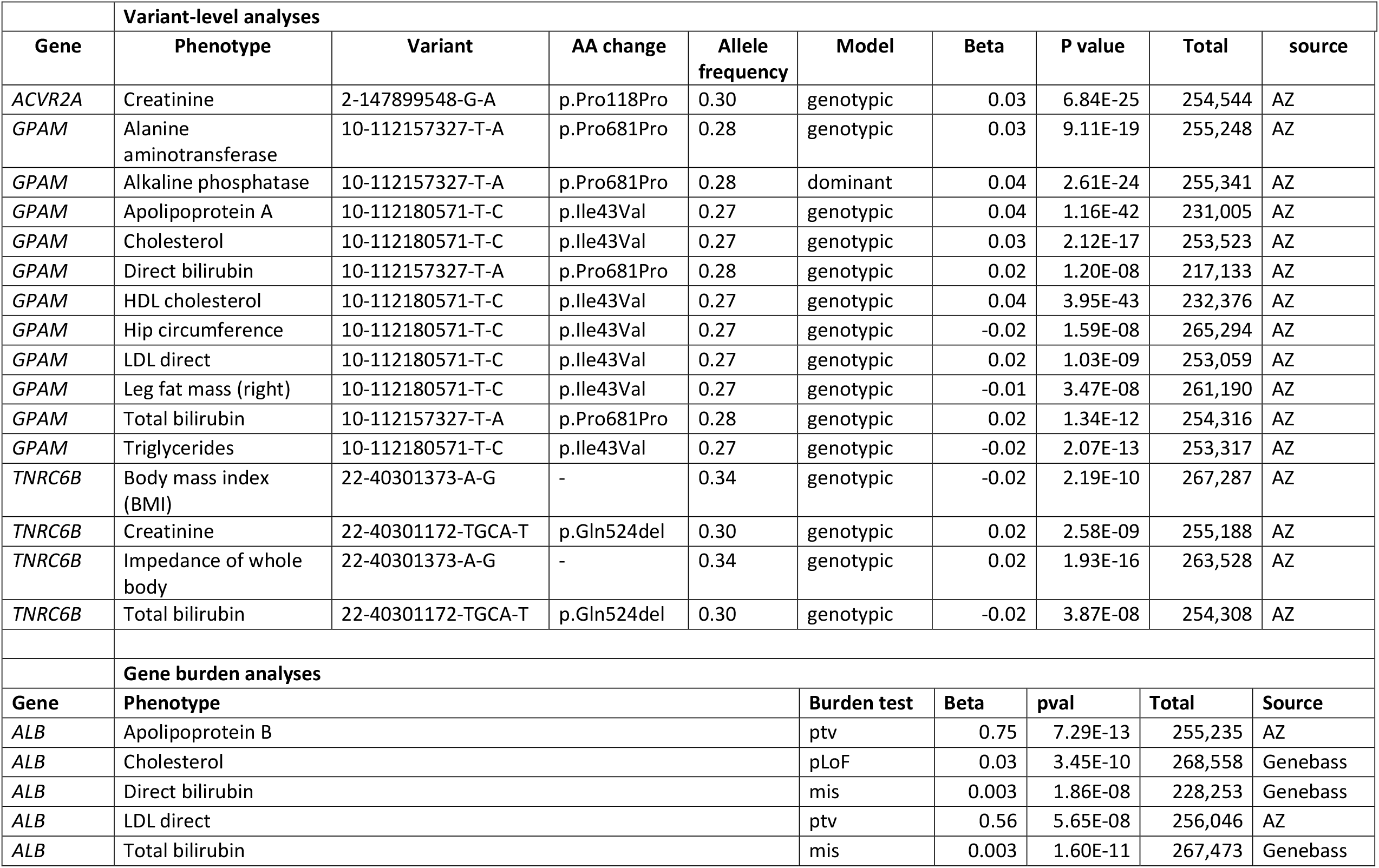
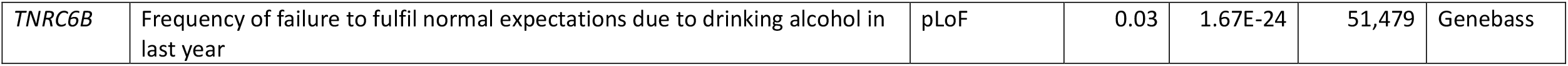
Top associations from rare variant and gene-burden analyses. Summary statistics were obtained from analyses of individual rare variants (using UK BioBank 300k Exomes) or gene-burden testing for pLoF or missense variants. AA, amino acid; AZ, Astrazeneca PheWAS; mis, missense variants; pLoF, predicted loss of function; ptv, protein truncating variant; snv, single nucleotide variant.

|  | Variant-level analyses |  |  |  |  |  |  |  |  |
| --- | --- | --- | --- | --- | --- | --- | --- | --- | --- |
| Gene | Phenotype | Variant | AA change | Allele frequency | Model | Beta | P value | Total | source |
| *ACVR2A* | Creatinine | 2-147899548-G-A | p.Pro118Pro | 0.30 | genotypic | 0.03 | 6.84E-25 | 254,544 | AZ |
| *GPAM* | Alanine aminotransferase | 10-112157327-T-A | p.Pro681Pro | 0.28 | genotypic | 0.03 | 9.11E-19 | 255,248 | AZ |
| *GPAM* | Alkaline phosphatase | 10-112157327-T-A | p.Pro681Pro | 0.28 | dominant | 0.04 | 2.61E-24 | 255,341 | AZ |
| *GPAM* | Apolipoprotein A | 10-112180571-T-C | p.Ile43Val | 0.27 | genotypic | 0.04 | 1.16E-42 | 231,005 | AZ |
| *GPAM* | Cholesterol | 10-112180571-T-C | p.Ile43Val | 0.27 | genotypic | 0.03 | 2.12E-17 | 253,523 | AZ |
| *GPAM* | Direct bilirubin | 10-112157327-T-A | p.Pro681Pro | 0.28 | genotypic | 0.02 | 1.20E-08 | 217,133 | AZ |
| *GPAM* | HDL cholesterol | 10-112180571-T-C | p.Ile43Val | 0.27 | genotypic | 0.04 | 3.95E-43 | 232,376 | AZ |
| *GPAM* | Hip circumference | 10-112180571-T-C | p.Ile43Val | 0.27 | genotypic | -0.02 | 1.59E-08 | 265,294 | AZ |
| *GPAM* | LDL direct | 10-112180571-T-C | p.Ile43Val | 0.27 | genotypic | 0.02 | 1.03E-09 | 253,059 | AZ |
| *GPAM* | Leg fat mass (right) | 10-112180571-T-C | p.Ile43Val | 0.27 | genotypic | -0.01 | 3.47E-08 | 261,190 | AZ |
| *GPAM* | Total bilirubin | 10-112157327-T-A | p.Pro681Pro | 0.28 | genotypic | 0.02 | 1.34E-12 | 254,316 | AZ |
| *GPAM* | Triglycerides | 10-112180571-T-C | p.Ile43Val | 0.27 | genotypic | -0.02 | 2.07E-13 | 253,317 | AZ |
| *TNRC6B* | Body mass index (BMI) | 22-40301373-A-G | - | 0.34 | genotypic | -0.02 | 2.19E-10 | 267,287 | AZ |
| *TNRC6B* | Creatinine | 22-40301172-TGCA-T | p.Gln524del | 0.30 | genotypic | 0.02 | 2.58E-09 | 255,188 | AZ |
| *TNRC6B* | Impedance of whole body | 22-40301373-A-G | - | 0.34 | genotypic | 0.02 | 1.93E-16 | 263,528 | AZ |
| *TNRC6B* | Total bilirubin | 22-40301172-TGCA-T | p.Gln524del | 0.30 | genotypic | -0.02 | 3.87E-08 | 254,308 | AZ |
|  | Gene burden analyses |  |  |  |  |  |  |  |  |
| Gene | Phenotype |  |  |  | Burden test | Beta | pval | Total | Source |
| *ALB* | Apolipoprotein B |  |  |  | ptv | 0.75 | 7.29E-13 | 255,235 | AZ |
| *ALB* | Cholesterol |  |  |  | pLoF | 0.03 | 3.45E-10 | 268,558 | Genebass |
| *ALB* | Direct bilirubin |  |  |  | mis | 0.003 | 1.86E-08 | 228,253 | Genebass |
| *ALB* | LDL direct |  |  |  | ptv | 0.56 | 5.65E-08 | 256,046 | AZ |
| *ALB* | Total bilirubin |  |  |  | mis | 0.003 | 1.60E-11 | 267,473 | Genebass |

**Table 2: Top associations from rare variant and gene-burden analyses.**
|  |  |  |  |  |  |  |
| --- | --- | --- | --- | --- | --- | --- |
| *TNRC6B* | Frequency of failure to fulfil normal expectations due to drinking alcohol in last year | pLoF | 0.03 | 1.67E-24 | 51,479 | Genebass |

### Germline variation at *GPAM*, but not other somatically mutated genes, is associated with liver phenotypes

We next explored whether more common germline variants might be associated with liver phenotypes in previously published datasets. We utilised the summary statistics from the UK BioBank cohort of 420,000 subjects[29] and searched for variants within these seven regions (**Figures 1-2, Table 3, & Supplementary Figure 1**). As previously described[16,17], we observed genome-wide significant variants within *GPAM*, associated with elevated serum levels of ALT (Table 2, e.g. rs10787429 C>T beta=.006, p=2.8×10^-39^), AST, and liver fat by MR imaging (e.g. rs11446981 T>TA beta=-.003, p=3.6×10^-13^). There were no significant associations between SNPs near any of the other recurrently mutated coding genes and disease correlates in NAFLD or ARLD.

**Figure 1:**
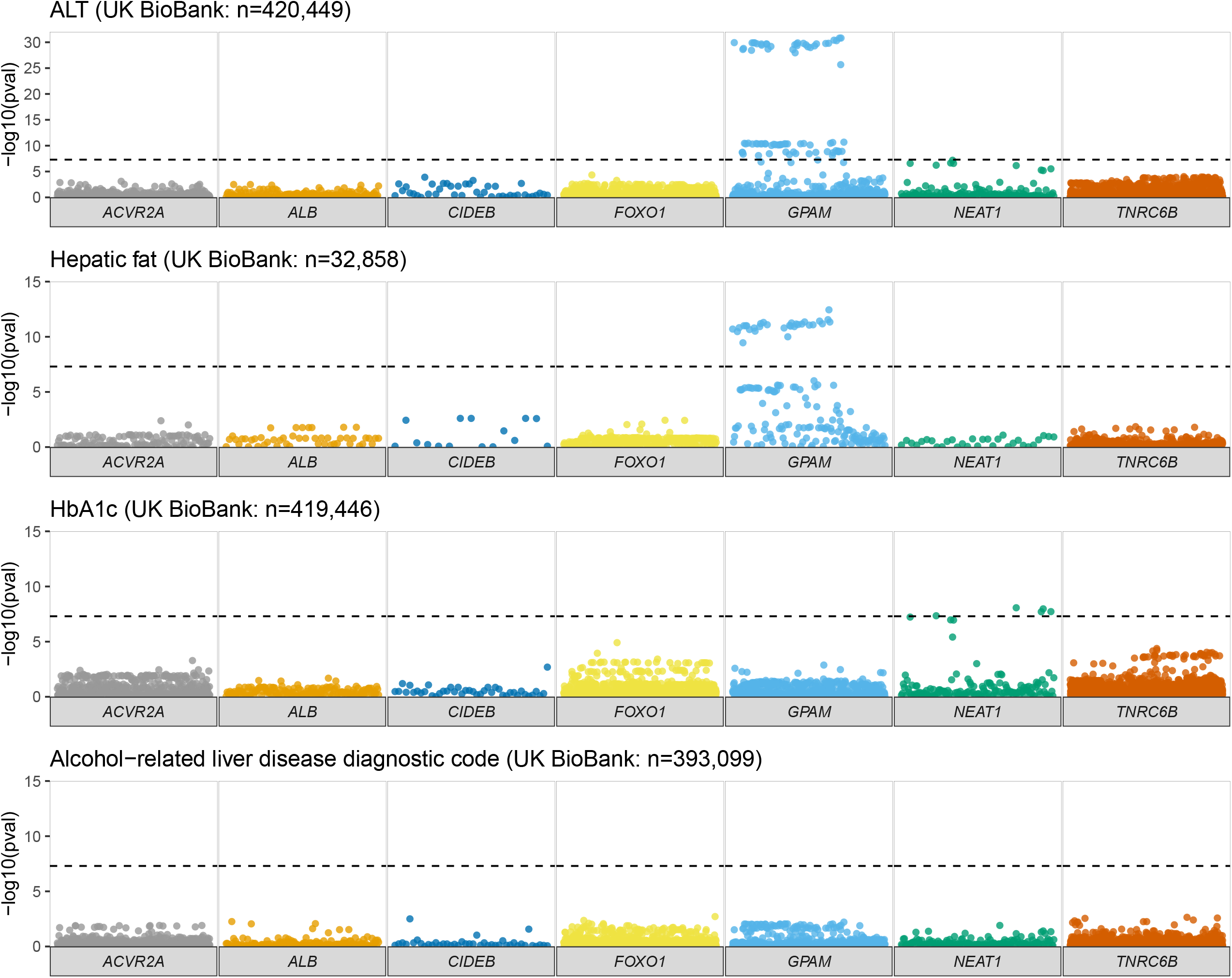
Association between common variants at recurrently mutated regions and markers of liver disease or glycaemic control. Manhattan plots focusing on the six protein-coding genes and one lncRNA (*NEAT1*) of interest, illustrating all variants within their genomic coordinates. -log10 p-value was obtained from summary statistics from the UK BioBank for alanine aminotransferase (ALT), liver fat (from Liu et al., 2021), glycosylated haemoglobin (HbA1c), and alcohol-related liver disease (ARLD). Significance was defined as p < 5×10^-8^.

**Table 3:** Genome-wide significant associations of common variants within regions of interest with eQTLs. Lead variants from the six protein-coding genes and one lncRNA (*NEAT1*) of interest associated with markers of liver disease or glycaemic control. GWAS summary statistics were obtained from the UK BioBank or Pazoki et al. (2021). Expression quantitative trait locus (eQTL) data were obtained from FIVEx. EAF, effect allele frequency.

| Variant | Gene | Trait | Beta | P value | EAF | Source | n | eQTL gene (tissue) | eQTL beta (p-value) | eQTL_study |
| --- | --- | --- | --- | --- | --- | --- | --- | --- | --- | --- |
| rs10787429 C > T | *GPAM* | ALT | 0.006 | 2.80E-39 | 0.27 | European GWAS meta-analysis | 753,010 | Nil |  |  |
| rs7096937 T > C | *GPAM* | AST | -0.02 | 2.77E-09 | 0.73 | UK BioBank | 419,034 | Nil |  |  |
| rs11446981 T > TA | *GPAM* | Liver fat | -0.06 | 3.60E-13 | 0.70 | UK BioBank | 32,858 | Nil |  |  |
| rs595366 T > A | *NEAT1* | ALT | -0.003 | 1.90E-11 | 0.27 | European GWAS meta-analysis | 753,010 | NEAT1 (adipose) | -0.4 (p=6.5x10<sup>-17</sup>) | FUSION |
| rs34743766 C > CA | *NEAT1* | HbA1c | 0.01 | 8.49E-09 | 0.17 | UK BioBank | 419,446 | NEAT1 (adipose) | 0.27 (p=9.3x10<sup>-17</sup>) | TwinsUK |

We validated these associations across a European-ancestry GWAS meta-analysis (n=753,010)[34] and in the BioBank Japan cohort (n=134,182[33]; **Figure 2, Supplementary Figure 1** & **Supplementary Table 5**). The magnitude of effect size for the lead variants within *GPAM* was similar to that for well-established loci in *HSD17B13* and *MTARC1*, but smaller than observed for risk variants in *PNPLA3* and *TM6SF2* (**Supplementary Table 6**). For example, for change in hepatic fat[31]: 10-113950257-T-TA in *GPAM* beta = -.06, p=3.60×10^-13^; compared to rs738409C>G in *PNPLA3*: beta = 0.22, p=1.5×10^-133^; and rs58542926T>C in *TM6SF2* beta = 0.33, p=2.5×10^-116^. Therefore, the only somatically-mutated gene for which germline variation was associated with liver disease was *GPAM*.

**Figure 2:**
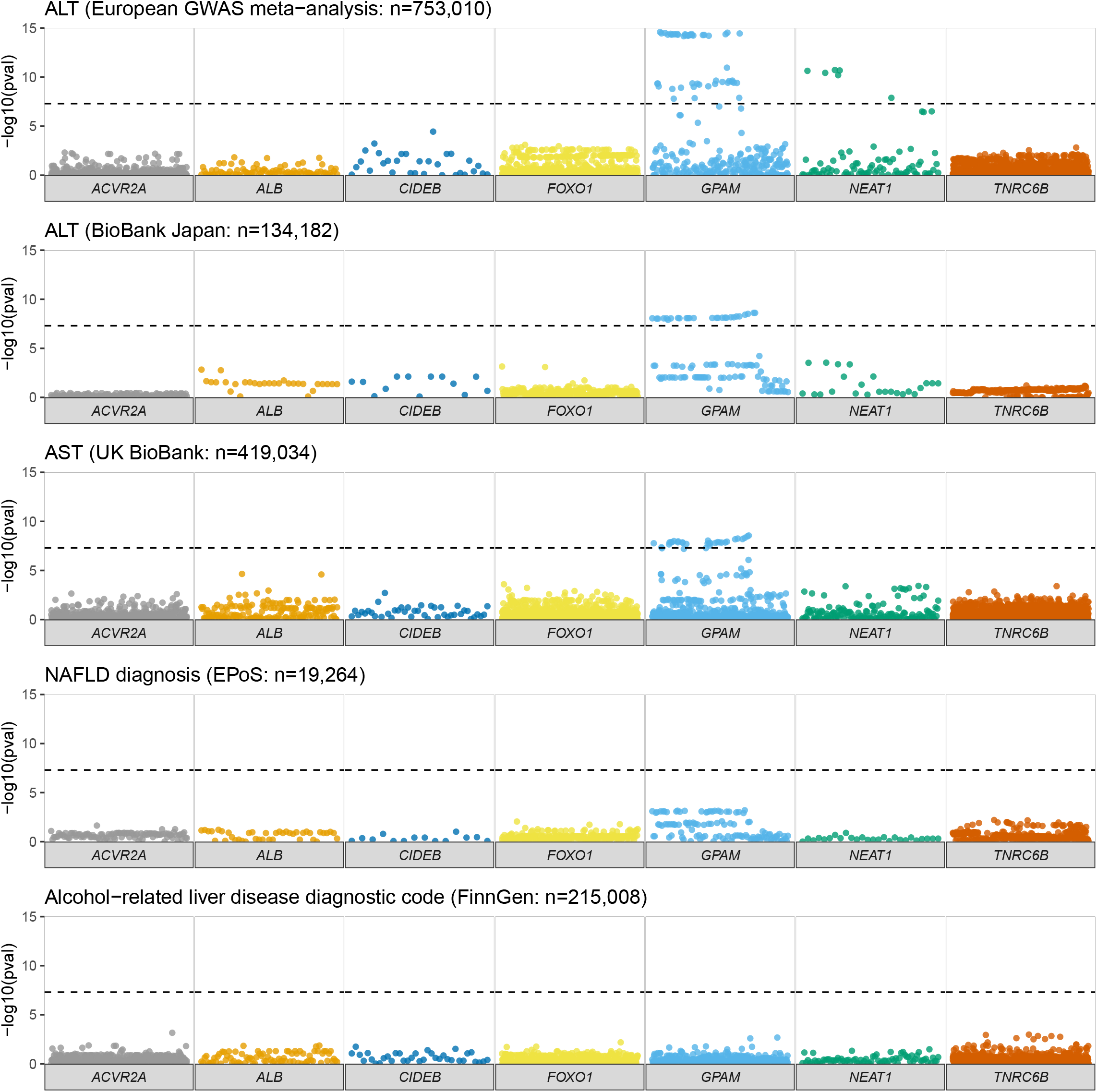
Association between common variants at recurrently mutated regions and markers of liver disease or glycaemic control. Manhattan plots focusing on the six protein-coding genes and one lncRNA (*NEAT1*) of interest, illustrating all variants within their genomic coordinates. -log10 p-value was obtained from summary statistics from the UK BioBank for aspartate aminotransferase (AST), hepatic fibrosis, and cirrhosis. Data on diagnosis of NAFLD was obtained from Anstee et al. (2021).

### Germline variation at the long non-coding RNA *NEAT1* is associated with liver phenotypes and glycaemic control

In the UK BioBank cohort we observed genome-wide significant variants within the lncRNA *NEAT1* for elevated serum ALT (**Figure 1**). This was replicated in the European ancestry ALT meta-analysis, though not in the BioBank Japan ALT results (**Figure 2**). The lead variant for *NEAT1* (rs595366 T>A) was also found to have an eQTL for *NEAT1* in both adipose and liver tissue.

However, no associations were identified between variants at *NEAT1* and categorical definitions of liver disease (e.g. diagnosis of NAFLD or ARLD, **Figures 1-2**) or severity of liver disease (e.g. cirrhosis). This was consistent across data from the UK BioBank and FinnGen (**Supplementary Figure 1 and Supplementary Table 5**) datasets.

Given the causal associations between insulin resistance and hepatic steatosis, we investigated whether variants were associated with HbA1c or a diagnosis of type 2 diabetes mellitus (T2DM). Variants in *NEAT1* were associated with HbA1c levels in the UK BioBank (e.g. rs34743766 C>CA, beta=0.1, p=8.5×10^-9^) and had an associated eQTL for *NEAT1* in adipose tissue, but this was not replicated for diagnosis of T2DM or other analyses of HbA1c (**Supplementary Figure 1**). These associations are interesting as expression levels of *NEAT1* have previously been associated with both the presence of diabetes[43] and the progression of diabetic complications[43,44].

### Germline variation at recurrently mutated genes is associated with metabolic phenotypes

We then explored whether common variants in or near these regions were associated with other related metabolic traits that affect patients with NAFLD, using a gene- (or region-) based phenome-WAS from two sources (Phenoscanner[25] and Common Metabolic Disease Portal[26], n=7,637-1,546,260, **Supplementary Table 7**). In addition to its association with ALT, variants in or near *GPAM* influenced the concentration of many serum lipids (**Table 4**), including LDL cholesterol and triglyceride levels. We also identified variants near *NEAT1* that were associated with a diagnosis of T2DM, waist-to-hip ratio and had a significant eQTL for *NEAT1* in adipose tissue. In addition, variants in or near *TNRC6B* were associated with BMI and creatinine, as were those in *ACVR2A*. Therefore, germline variation in these genes may impact on related phenotypes that accompany the metabolic syndrome.

**Table 4:** Associations between common variants in or near regions of interest and related metabolic traits. Gene-based PheWAS performed using Phenoscanner and Common Metabolic Disease Portal to identify common variants significantly associated with the six protein-coding genes and one lncRNA (*NEAT1*) of interest. eQTL column provides the gene that have significant eQTLs for that variant, the tissues, and the direction (+, positive eQTL; -, negative eQTL). eQTL data were obtained using the Qtlizer package for R.

| Gene | Variant | Trait | Beta (SE) | P value | n | eQTL |
| --- | --- | --- | --- | --- | --- | --- |
| ACVR2A | rs13008838 A>G | eGFR-creat (serum creatinine) | -0.003 (0.0004) | 4.97E-15 | 754,661 | ACVR2A: Adipose - Subcutaneous (+), Muscle skeletal (+), Artery - Tibial (+), Artery - Aorta (+), Pancreas (+), Artery - Coronary (+), Adipose - Visceral (Omentum) (+) |
| ACVR2A | rs3764955 G>C | Serum creatinine | 0.020 (0.003) | 2.01E-08 | 182,901 | ACVR2A: Artery - Coronary (+), Adipose - Visceral (Omentum) (+), Adipose - Subcutaneous (+), Pancreas (+), Artery - Aorta (+), Artery - Tibial (+), Muscle skeletal (+) |
| CIDEB | rs12590407 G>A | Diastolic blood pressure | -0.010 (0.002) | 9.11E-12 | 552,754 |  |
| FOXO1 | rs2253001 A>T | Basal metabolic rate | 0.010 (0.002) | 1.03E-08 | 331,307 |  |
| GPAM | rs10787429 T>C | Alanine transaminase | -0.030 (0.005) | 3.44E-09 | 141,341 | GPAM: Artery - Tibial (-) |
| GPAM | rs7898213 T>C | Alkaline phosphatase | -0.030 (0.004) | 5.00E-11 | 112,189 |  |
| GPAM | rs2792759 C>T | Bilirubin | -0.010 (0.001) | 4.10E-09 | 467,109 | GPAM: Artery - Tibial (-) |
| GPAM | rs2297991 T>C | HDL cholesterol | -0.030 (0.003) | 5.21E-20 | 191,159 | GPAM: Artery - Tibial (-) |
| GPAM | rs1129555 A>G | LDL cholesterol | 0.030 (0.004) | 1.00E-15 | - | GPAM: Artery - Tibial (-) |
| GPAM | rs1129555 A>G | Total cholesterol | 0.030 (0.004) | 2.00E-18 | - | GPAM: Artery - Tibial (-) |
| GPAM | rs2254537 T>A | Triglycerides | 0.020 (0.002) | 8.23E-17 | 482,392 | GPAM: Artery - Tibial (-) |
| NEAT1 | rs10896037 A>G | Chronic kidney disease | 0.060 (0.009) | 5.98E-10 | 140,966 | NEAT1: Adipose - Visceral (Omentum) (-), Muscle skeletal (-), Adipose - Subcutaneous (-), Artery - Aorta (-) |
| NEAT1 | rs12801636 G>A | Coronary artery disease | -0.040 (0.004) | 6.74E-17 | 1,546,260 |  |
| NEAT1 | rs2306363 G>T | Diastolic blood pressure | -0.030 (0.002) | 1.39E-28 | 552,754 |  |
| NEAT1 | rs4930319 G>C | eGFR-creat (serum creatinine) | 0.003 (0.0004) | 8.69E-27 | 757,454 | NEAT1: Adipose - Subcutaneous (-), Adipose - Visceral (Omentum) (-), Artery - Aorta (-), Muscle skeletal (-) |
| NEAT1 | rs2236682 G>T | Serum creatinine | 0.020 (0.004) | 2.66E-10 | 182,901 | NEAT1: Muscle skeletal (-), Artery - Aorta (-), Adipose - Visceral (Omentum) (-), Adipose - Subcutaneous (-) |
| NEAT1 | rs2306363 G>T | Systolic blood pressure | -0.020 (0.002) | 3.90E-20 | 550,853 |  |
| NEAT1 | rs10750766 C>A | Triglycerides | 0.020 (0.002) | 9.96E-15 | 459,761 |  |
| NEAT1 | rs947791 G>A | Type 2 diabetes | 0.050 (0.004) | 2.71E-16 | 1,016,100 | NEAT1: Artery - Aorta (+), Pancreas (+), Muscle skeletal (+), Artery - Tibial (+), Adipose - Visceral (Omentum) (+), Adipose - Subcutaneous (+) |
| NEAT1 | rs11227217 C>T | Waist-hip ratio | 0.020 (0.002) | 2.60E-15 | 997,776 | NEAT1: Adipose - Visceral (Omentum) (+), Muscle skeletal (+), Adipose - Subcutaneous (+), Artery - Aorta (+), Artery - Tibial (+), Pancreas (+) |
| NEAT1 | rs4645917 G>A | Waist-hip ratio adj BMI | -0.030 (0.002) | 5.05E-18 | 969,126 | NEAT1: Muscle skeletal (-), Artery - Tibial (-) |
| TNCR6B | rs4820410 A>G | BMI | -0.020 (0.001) | 1.72E-26 | 1,326,100 | TNRC6B: Artery - Tibial (+) |
| *TNCR6B* | rs2294352 G>A | eGFR-creat (serum creatinine) | 0.010 (0.001) | 2.90E-32 | 772,751 |  |
| *TNCR6B* | rs470113 A>G | Pulse pressure | 0.020 (0.003) | 4.33E-08 | 317,539 | TNRC6B: Artery - Aorta (+), Artery - Tibial (+), Artery - Aorta (+), Artery - Tibial (+) |
| *TNCR6B* | rs2294352 G>A | Serum creatinine | -0.030 (0.004) | 1.06E-15 | 191,380 |  |
| *TNRC6B* | rs5995840 T>C | Body mass index | 0.030 (0.004) | 1.02E-11 | 173,430 | TNRC6B: Artery - Aorta (+), Artery - Tibial (+), Atherosclerotic aortic root NA |

## Discussion

Investigating the genetics of fatty liver disease has the potential to inform our understanding of disease biology and facilitate clinical risk stratification for affected patients[3]. We recently identified six protein-coding genes and one lncRNA (*NEAT1*) that are enriched for somatic mutations in patients with NAFLD or ARLD[11]. In this study we find that germline variation in only one, *GPAM*, was robustly associated with markers of liver disease. Therefore, genetic risk scores capturing only germline variation will not include the pathogenicity conveyed by these regions and additional strategies may be needed to understand the prognostic implications of these somatic mutations.

For five (*ACVR2A, ALB, CIDEB, FOXO1, TNRC6B*) of the six protein-coding genes under investigation in this study, we found no evidence for association between germline variation and liver disease. These observations were consistent across multiple data sources, traits, genetic ancestries, and analysis methodologies, for example both common variant analyses and rare-variant gene burden testing. This was an unexpected observation, given the genetic evidence for their selective advantage in hepatocyte clones in diseased non-malignant NAFLD and ARLD[11]. Moreover, *CIDEB* and *FOXO1* have well-established functions as a lipid droplet-associated protein[12] and a component of the insulin signalling cascade[14], respectively. Conversely, our previous study did not identify acquired somatic mutations in genes with strong germline associations with liver disease (e.g. *PNPLA3, TM6SF2*). Collectively, this suggests that the influence of germline and acquired variants on parenchymal liver disease occurs through independent mechanisms (and genes).

It should be noted that the methodology employed in this study does not exclude the possibility that individuals with rare loss of function mutations in these genes may have liver-related phenotypes. Identification and studying human knock-outs for these genes is an alternative strategy for investigating whether germline variation plays a role in liver disease, as has been illustrated for other conditions[45,46]. However, our data suggest that these individuals will be very rare.

Our analysis has replicated the known associations between variants in or near *GPAM* and liver fat, as well as ALT and AST. Whilst we did not show these variants to influence diagnosis of NAFLD, this has been demonstrated by others[18] with larger sample sizes, variants in *GPAM* (particularly p.Ile43Val) are associated with radiological and histological diagnosis of NAFLD[16–19]. However, It is important to note that variants in *GPAM* have a comparatively small effect size compared to variants in *PNPLA3* and *TM6SF2*, but similar to those in *HSD17B13* and *MTARC1*.

We identified borderline associations between variants in *NEAT1*, a long non-coding RNA (lncRNA), with ALT and HbA1c. Our more broad analyses implicated *NEAT1* in influencing multiple metabolic traits (e.g. serum triglycerides, diagnosis of T2DM, and coronary artery disease), that are of potential relevance to patients with NAFLD. These results point towards a primary role on insulin resistance, potentially through modulation of adipose tissue biology, as several variants in this lncRNA also had significant eQTLs in adipose tissue. The biology of *NEAT1* is poorly understood, but there is some *in vitro* evidence for its role in adipogenesis[47]. We suggest that the subtle effects of germline variants in *NEAT1* on ALT are likely indirect, via perturbation of insulin resistance and/or development of T2DM[48], however further work is required to establish this. This data also underlines the principle that the genomic regions enriched for somatic mutations in our original analysis are principally those involved in metabolism.

Clinically, one aim of human genetics is to stratify patients into high- and low- risk groups for disease progression using polygenic gene scores. Such an approach can identify individuals with a five-fold increased risk of coronary artery disease[49]. This would be of particular use for NAFLD and ARLD, both common conditions where only a minority of individuals progress to liver-related clinical events. To date there have been four PGS published for liver disease[8,9,50,51], all derived using genome-wide significant hits, and therefore none of our seven genomic regions of interest were included. If a genome-wide PGS were derived[52], which included weighting from sub-genome wide-significant variants, then variants in or near *GPAM* would contribute. However, they would still receive comparatively minimal weighting compared to variants in *PNPLA3* and *TM6SF2*. More broadly, it is not clear how the magnitude of prognostic implication would compare for germline variation risk scores compared to somatic mutations, as the prognostic implication of these remains unknown. Our results illustrate that integration of somatic mutants into prognostic tools will be a complex process and separate from existing methods for polygenic gene scores.

To understand the role that somatically mutated genes could play in germline PGS there is a need for more independent cohorts with liver-directed exomic or targeted sequencing. This will facilitate PGS derivation and validation against hard clinical outcomes (e.g. liver transplantation) to formally test whether variants in or near *ACVR2A, ALB, CIDEB, FOXO1*, and *TCNR6B* are prognostic.

## Conclusion

Out of seven genomic regions with strong evidence for playing a role in NAFLD and ARLD through somatic mutations, only germline variation in *GPAM* is associated with liver disease. Other genes analysed, including the lncRNA *NEAT1*, have a role in metabolic pathways Therefore, genes with pathogenic somatic mutations in NAFLD and ARLD are distinct to those that confer germline risk and would not be captured by polygenic risk scores. Novel approaches will be required to integrate somatic and germline variation with clinical variables for risk prediction algorithms.

## Data Availability

All summary statistics are available in the article and supplementary data.
Code used in analysis is available via https://doi.org/10.5281/zenodo.4656979.

## Acknowledgements

We want to acknowledge the participants and investigators of FinnGen, UK BioBank, BioBank Japan, EPoS, Million Veterans, and MAGIC Consortium studies.

## Figure and table legends

**SupTable 1: Summary of data sources for genome-wide association study summary statistics used, including number of participants.**

**SupTable 2: List of all included somatic variants with full annotation from Variant Effect Predictor.**

**SupTable 3: Associations between somatic variants previously identified in the germline and metabolic traits from the Common Metabolic Disease Portal. Critical p-value for significance p<5.2×10^-4^.**

**SupTable 4: Full results from rare exonic variant and gene-burden analyses.**

**SupTable 5: Full results from single common variant analysis for markers of liver disease from all studies (discovery and replication) across all regions of interest.**

**SupTable 6: Comparison of lead variants within GPAM region with well-established genome-wide significant variants across all studied traits.**

**SupTable 7: Full results from gene-based PheWAS for common variants for associated metabolic traits from Phenoscanner and Common Metabolic Disease Knowledge Portal.**

**SupFig1:**
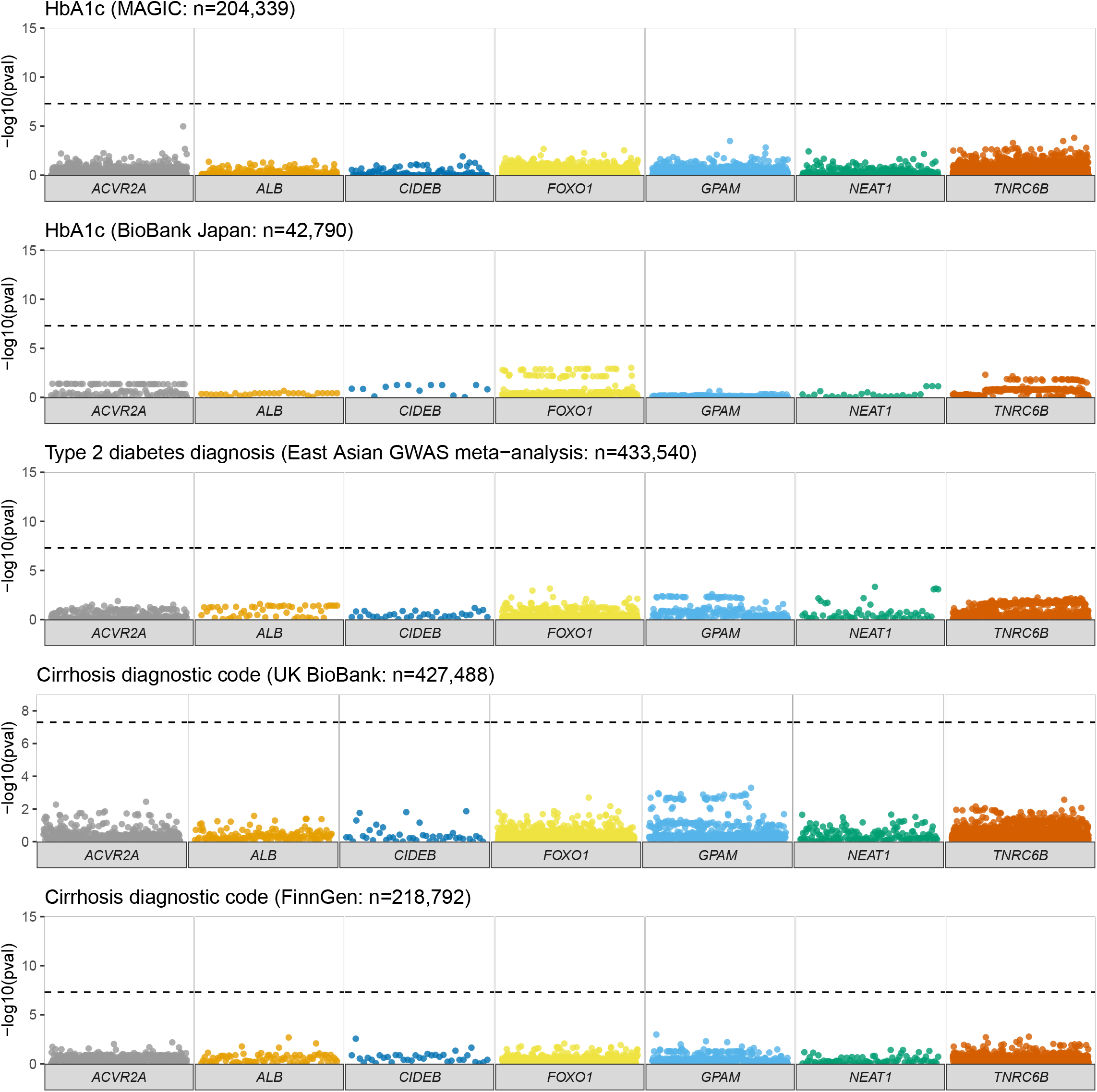
Replication analyses for association between common variants and markers of liver disease or glycaemic control. Manhattan plot focusing on the six protein-coding genes and one lncRNA (*NEAT1*) of interest, illustrating all variants within their genomic coordinates. - log10 p-value was obtained from summary statistics from Paroki et al. (2021) and BioBank Japan for ALT, Spracklen et al. (2020) for diagnosis of type 2 diabetes mellitus (T2DM), and from FinnGen for diagnosis of alcohol-related liver disease and cirrhosis.

## Notes

**Funding:** JPM is supported by a Wellcome Trust fellowship (216329/Z/19/Z); MH is supported by a CRUK Advanced Clinician Scientist fellowship (C52489/A19924); CRUK-OHSU Project Award (C52489/A29681) and CRUK Accelerator award to the HUNTER consortium (C18873/A26813).

### Competing Interest Statement

MH is a co-inventor on a patent detailing the finding of recurrent somatic mutations in chronic liver disease.

### Funding Statement

JPM is supported by a Wellcome Trust fellowship (216329/Z/19/Z); MH is supported by a CRUK Advanced Clinician Scientist fellowship (C52489/A19924); CRUK-OHSU Project Award (C52489/A29681) and CRUK Accelerator award to the HUNTER consortium (C18873/A26813).

### Author Declarations

UK BioBank Summary Stats: https://pan.ukbb.broadinstitute.org/ FinnGen: https://www.finngen.fi/en BioBank Japan: http://jenger.riken.jp/en/ Other GWAS Summary Statistics: https://www.ebi.ac.uk/gwas/downloads/summary-statistics Common Metabolic Disease Knowledge Portal: https://hugeamp.org/ MAGIC summary stats: https://magicinvestigators.org/downloads/

